# Nigral volume as a marker for Lewy body copathology in Alzheimer’s disease

**DOI:** 10.64898/2026.11.23.26368759

**Authors:** Jason Langley, Kristy S. Hwang, Xiaoping P. Hu, the Alzheimer’s Disease Neuroimaging Initiative

## Abstract

**Background:** Lewy body (LB) copathology in Alzheimer’s disease (AD) is associated with earlier symptom onset and faster cognitive decline. Depigmentation of substantia nigra pars compacta (SNc) volume is seen in both Parkinson’s disease and dementia with Lewy bodies. We examined whether lower antemortem SNc volume was associated with autopsy confirmed LB pathology in patients with AD or mild cognitive impairment (MCI).

**Methods:** Seventy-three participants with a clinical diagnosis of AD or MCI, neuromelanin-sensitive MRI, and subsequent neuropathologic assessment from the Alzheimer’s Disease Neuroimaging Initiative were studied. At autopsy, 33 participants had LB pathology (LB+) and 40 had no LB pathology (LB−). We compared SNc volume between groups using analysis of covariance adjusted for age and sex. Within the LB+ group, we assessed the association between SNc volume and nigral hypopigmentation using Spearman correlation. In a secondary analysis, we tested whether adding SNc volume to a logistic regression mode containing age, sex, and executive function improved model fit for autopsy-defined LB status.

**Results:** LB+ participants had greater nigral hypopigmentation at autopsy and lower antemortem SNc volume than LB− participants. The adjusted mean difference in SNc volume (LB+ minus LB−) was −66 mm3 (95% confidence interval −115.9 to −16.2, P=0.010). Within the LB+ group, lower SNc volume was associated with greater hypopigmentation (Spearman ρ=−0.373, P=0.032). Among 71 participants with complete model data, adding SNc volume improved model fit (likelihood ratio χ2(1)=8.67, P=0.003). The combined model had an in sample area under the receiver operating characteristic curve of 0.770 (95% confidence interval: 0.661-0.879).

**Conclusion:** The correspondence between lower antemortem SNc volume and greater hypopigmentation at autopsy supports the use of neuromelanin-sensitive MRI to characterize nigral involvement in AD or MCI with LB copathology. These findings are based on a single cohort, and the interval of several years between imaging and autopsy limits how closely the pigmentation ratings reflect nigral integrity at imaging.

## 1. Introduction

Alzheimer’s disease (AD) is the most common neurodegenerative dementia ^1^ and is characterized neuropathologically by extracellular β-amyloid (Aβ) plaques and neurofibrillary tangles containing hyperphosphorylated tau ^2-4^. Mild cognitive impairment (MCI) involves cognitive decline that does not meet the criteria for dementia, and those with MCI and underlying AD pathology are at an increased risk of progression to AD dementia ^5^. Autopsy studies have found that up to 60% of AD cases have Lewy body (LB) pathology ^6-8^. In AD, LB copathology is associated with more rapid cognitive decline ^9^, parkinsonism ^10^, younger symptom onset ^11^, and shorter survival times ^12^.

LB copathology in AD is associated with greater visuospatial and executive function impairment ^9,13^. Its presence can be difficult to identify in AD patients because cognitive assessment does not reliably distinguish AD with concomitant LB pathology from AD alone ^14,15^. Cerebrospinal fluid-based α-synuclein amplification assays (SAAs) enable antemortem detection of α-synuclein seeding activity and show substantial agreement with autopsy-confirmed LB pathology ^16^. These assays provide evidence of α-synuclein pathology, but do not directly characterize the integrity of the affected brain structures. Complementary measures of regional brain neurodegeneration may help characterize the consequences of LB copathology in AD patients.

Postmortem studies have demonstrated greater neuronal loss and depigmentation in the substantia nigra of AD patients with LB copathology than in those with pure AD pathology ^10,17^. Neuromelanin-sensitive MRI can visualize neurons in substantia nigra pars compacta (SNc) using incidental or explicit magnetization transfer (MT) effects ^18-21^. Nigral volume derived from neuromelanin-sensitive MRI is reduced in overt ^22-27^ and prodromal ^28,29^ Parkinson’s disease, while reduced volume has been reported in dementia with Lewy bodies ^30^. These observations suggest that neuromelanin-sensitive MRI may also detect nigral changes associated with LB copathology in individuals with AD or MCI.

## 2. Methods

### 2.1 ADNI Overview

Data used in this study were obtained from the ADNI database (adni.loni.usc.edu). The ADNI was launched in 2003 as a public-private partnership, led by Principal Investigator Michael W. Weiner, MD. Up-to-date information can be found at www.adni-info.org. The ADNI study was approved by the local Institutional Review Boards (IRBs) of all participating sites. Study subjects and, if applicable, their legal representatives, gave written informed consent at the time of enrollment for imaging data, genetic sample collection and clinical questionnaires.

### 2.2 Participants

The Alzheimer’s Disease Neuroimaging Initiative (ADNI) database (adni.loni.usc.edu) was queried for individuals with neuropathologic data, T1-weighted images, and neuromelanin-sensitive MRI images. Each participant’s final ADNI1 imaging visit was selected. Participants with a clinical diagnosis of MCI or AD at that imaging visit were included, yielding 40 participants without LB pathology (AD/MCI LB−) and 33 participants with LB pathology (AD/MCI LB+). Cognitive and clinical measures from the same visit included the Alzheimer’s Disease Assessment Scale cog-11 (ADAS11), mini-mental state examination (MMSE), clinical dementia rating (CDR), trail making test, and University of Washington (UW) composite scores (executive function, visuospatial, and memory). Imaging data were downloaded between March 2022 and October 2026.

### 2.3 Neuropathologic analysis

ADNI neuropathologic evaluations follow the National Institute on Aging and Alzheimer’s Association guidelines and Dementia with Lewy Bodies Consortium criteria. Nigral hypopigmentation was rated on a four-level scale: 0 – no loss, 1 – slight loss, 2 – moderate loss, 3 – extensive. Neurofibrillary tangle pathology was characterized using Braak stage. LB pathology was classified by its distribution as absent, brainstem-predominant, limbic (transitional), neocortical (diffuse), amygdala-predominant, or olfactory-only. Participants with LB pathology in any assessed region were classified as AD/MCI LB+ while participants without LB pathology were classified as AD/MCI LB−.

### 2.4 Synuclein amplification assays

Cerebrospinal fluid-based α-synuclein amplification assay (SAA) results were obtained from the ADNI database. Assays were performed by Amprion Clinical Laboratory using a clinically validated method, as previously described ^16^. Samples were tested in triplicate, and α-synuclein aggregation was monitored using thioflavin T fluorescence. Results indicated Parkinson’s disease or Dementia with Lewy Bodies-like seeding activity were classified as SAA positive, whereas those with no detectable seeding activity were classified as SAA negative. Indeterminate results were reported separately and excluded from calculations of concordance with autopsy-defined LB status.

### 2.5 Image processing

Neuromelanin-sensitive images were obtained from the first echo of an interleaved dual-echo turbo spin echo (TSE) acquisition, which contains incidental magnetization transfer effects ^31^. Acquisition parameters were echo time (TE) TE_1_/TE_2_/ repetition time (TR)=11/101/3270 ms, field of view (FOV)=240×213 mm^2^, voxel size=0.9×0.9×3 mm^3^, 48 slices. T_1_-weighted structural images in the ADNI1 cohort were used for registration to common space.

Registration to Montreal Neurological Institute (MNI) standard space was performed using fMRIB Software Library (FSL) ^32,33^. T_1_-weighted images were skull stripped using the brain extraction tool and registered to the 2 mm T_1_-weighted MNI template using an affine registration with flirt, followed by a nonlinear registration with fnirt. The second echo from the dual echo TSE sequence was brain extracted and registered to the brain extracted T_1_-weighted image using rigid body registration with a boundary-based cost function. The inverse of the T_1_ to MNI transform was concatenated with the T_1_-TSE transform and used to map the cerebral peduncle reference region and probabilistic SNc atlas from MNI space to native TSE space ^34^.

SNc segmentation was performed on the first echo from the TSE image using a thresholding method. A reference region in the cerebral peduncle was transformed into native TSE space as described above. Segmentation was restricted to the anatomical location of the SNc using the transformed atlas ^34^. Voxels with intensities exceeding the mean of the reference region plus 3 standard deviations were considered part of SNc.

### 2.6 Statistical Analysis

Group differences in sex distribution were assessed using Pearson’s chi-square test. Differences in continuous demographic and clinical measures were assessed using one-way analysis of variance. Group differences in SNc volume were assessed using analysis of covariance, adjusting for age at imaging and sex. Within the AD/MCI LB+ group, the association between SNc volume and nigral hypopigmentation was assessed using Spearman’s rank correlation. To assess the influence of individual participants on the association between SNc volume and nigral hypopigmentation, the Spearman correlation was recalculated after omitting each LB+ participant in turn. A partial Pearson correlation was used to assess this association after adjustment for the imaging to autopsy interval. Associations between SNc volume and cognitive measures were assessed using Pearson correlations and partial correlations adjusting for age and education.

Binary logistic regression was used to evaluate whether SNc volume provided additional information about LB status beyond age, sex, and executive dysfunction. An initial model included age, sex, and the UW executive function composite score. SNc volume was then added, and improvement in model fit was assessed using a likelihood-ratio test. Receiver operating characteristic (ROC) curves and areas under the curve (AUCs), with 95% confidence intervals, were calculated for SNc volume, executive function, and the combined model. Combined model discrimination was evaluated in the same sample used to fit the model.

Concordance between SAA classification and autopsy defined LB status was calculated among participants with determinate SAA results. Participants with missing data were excluded from analyses requiring those measures. One participant with an invalid imaging to autopsy interval was excluded from analyses involving that interval. All statistical tests were two-sided with statistical significance defined as P<0.05. All statistical analyses were performed using IBM SPSS Statistics software version 24 (IBM Corporation, Somers, NY, USA) and results are reported as mean ± standard deviation.

## 3. Results

### 3.1 Sample Demographics

Thirty-three AD/MCI LB+ and 40 AD/MCI LB− participants had autopsy and imaging data. Demographic data is summarized in Table 1. The groups did not differ significantly in sex distribution, age at imaging or autopsy, imaging to autopsy interval, Braak stage, education, MMSE, CDR global score, or ADAS Q4 score (Ps>0.108). Compared with AD/MCI LB− participants, AD/MCI LB+ participants had longer Trails B completion times (P=0.002), higher ADAS 11 scores (P=0.029), and lower University of Washington (UW) executive function (P=0.004), visuospatial (P=0.034), and memory (P=0.038) composite scores, indicating poorer cognitive performance.

**Table 1.** Demographic information for all participants in the AD/MCI groups with and without Lewy bodies used in imaging analyses. Data is presented as mean ± standard deviation unless noted otherwise. A Chi-square test was used to examine differences in gender. Two-tailed *t*-tests were used for group comparisons of all ages, education, MMSE, ADAS11, and Trails B from which *P* values are shown. Wilcoxon signed rank tests were used for group comparisons of nigral neuronal loss, Braak stage, and CDR Global. Amy – amygdala. AD – Alzheimer’s disease. BS – brainstem predominant. LB – Lewy bodies. Lim – Limbic (transitional). MCI – Mild Cognitive Impairment. MMSE – Mini-Mental State Exam. NC – Neocortical (diffuse). CDR – clinical dementia rating, ADAS – Alzheimer’s Disease Assessment Scale.

|  | AD/MCI LB-<br>*N*=40 | AD/MCI LB+<br>*N*=33 | Test<br>Statistic | P |
|---|---|---|---|---|
| Gender [Male/Female] | 26/14 | 27/6 | 2.571 | 0.109 |
| Age at imaging [years] | 80.0 ± 5.3 | 80.2 ± 7.5 | 0.020 | 0.887 |
| Age at autopsy [years] | 84.6 ± 6.5 | 83.9 ± 7.6 | 0.169 | 0.693 |
| Interval [years] | 5.0 ± 4.5 | 4.0 ± 3.7 | 1.048 | 0.309 |
| Nigral hypopigmentation | 0.76 ± 0.58 | 1.24 ± 0.61 | **12.152** | **<10<sup>-3</sup>** |
| LB Distribution [BS/Lim/NC/Amy] | None | 2/7/15/9 | N/A | N/A |
| Braak stage | 4.33 ± 1.44 | 4.18 ± 1.64 | 0.180 | 0.673 |
| Education [years] | 15.7 ± 2.7 | 15.9 ± 3.0 | 0.166 | 0.685 |
| CDR global | 0.76 ± 0.72 | 1.02 ± 0.81 | 2.113 | 0.150 |
| MMSE | 23.8 ± 6.3 | 21.4 ± 6.2 | 2.655 | 0.108 |
| ADAS11 | 15.19 ± 10.54 | 21.37 ± 13.24 | **4.995** | **0.029** |
| ADAS Q4 | 6.86 ± 3.42 | 7.61 ± 3.01 | 0.984 | 0.325 |
| Trails A [s] | 49.7 ± 32.1 | 69.9 ± 46.0 | **4.446** | **0.039** |
| Trails B [s] | 137.3 ± 89.4 | 210.5 ± 84.8 | **10.790** | **0.002** |
| UW EF1 | -0.15 ± 1.31 | -1.01 ± 1.13 | **8.651** | **0.004** |
| UW VS | -0.21 ± 1.07 | -0.78 ± 1.21 | **4.683** | **0.034** |
| UW Memory | -0.38 ± 1.31 | -1.00 ± 1.19 | **4.491** | **0.038** |

CSF α-synuclein seed amplification assay (SAA) results were available for 19 AD/MCI LB+ and 27 AD/MCI LB− participants. α-synuclein seeding activity was positive in 15 and negative in 3 LB+ participants. One assay was indeterminate. Among LB− participants, seeding activity was detected in one participant and was not detected in 26 participants. Among participants with determinate SAA results, SAA classification was concordant with autopsy defined LB status in 41 of 45 cases (91.1%). SAA results are summarized in Table 2.

**Table 2.** SAA positivity in the cohort.

|  | AD/MCI LB- | AD/MCI LB+ |
|---|---|---|
| SAA Positive | 1 | 15 |
| SAA Negative | 26 | 3 |
| SAA Indeterminate | 0 | 1 |
| Unavailable | 13 | 14 |

### 3.2 Nigral hypopigmentation and SNc volume

AD/MCI LB+ participants had higher nigral hypopigmentation scores than AD/MCI LB− participants (LB+: 1.24 ± 0.61; LB−: 0.76 ± 0.58; P<10-3), indicating greater nigral pigmentation loss. After adjustment for age and sex, SNc volume was lower in AD/MCI LB+ participants than in AD/MCI LB− participants (LB+ adjusted mean: 265.7 mm3 ± 113.3 mm3; LB− adjusted mean: 331.8 mm3 ± 101.3 mm3; F=6.978; P=0.010). The adjusted mean difference was −0.66 mm3 (95% confidence interval: −115 – −16.2). Age (F=0.954; P=0.332) and sex (F=3.755; P=0.057) were not significant covariates in the model. This comparison is shown in Figure 2.

**Figure 1.**
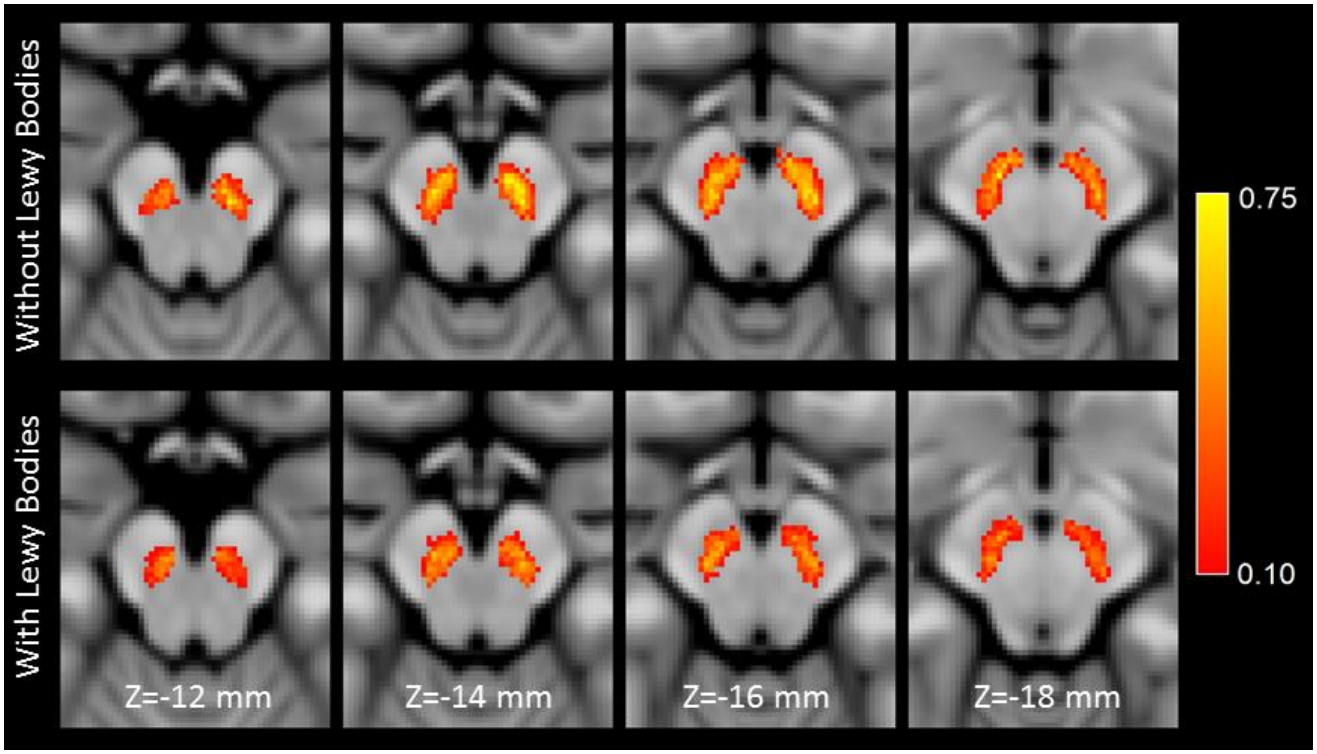
A comparison of SNc population atlases in AD/MCI LB−(top row) and AD/MCI LB+ (bottom row) groups. The AD/MCI LB+ group has reduced volume as compared to the AD/MCI LB− group in slices Z=−12 mm and Z=−14 mm. For each group, the SNc population atlas was created by transforming nigral volumes from individual participants to MNI space and then averaging.

**Figure 2.**
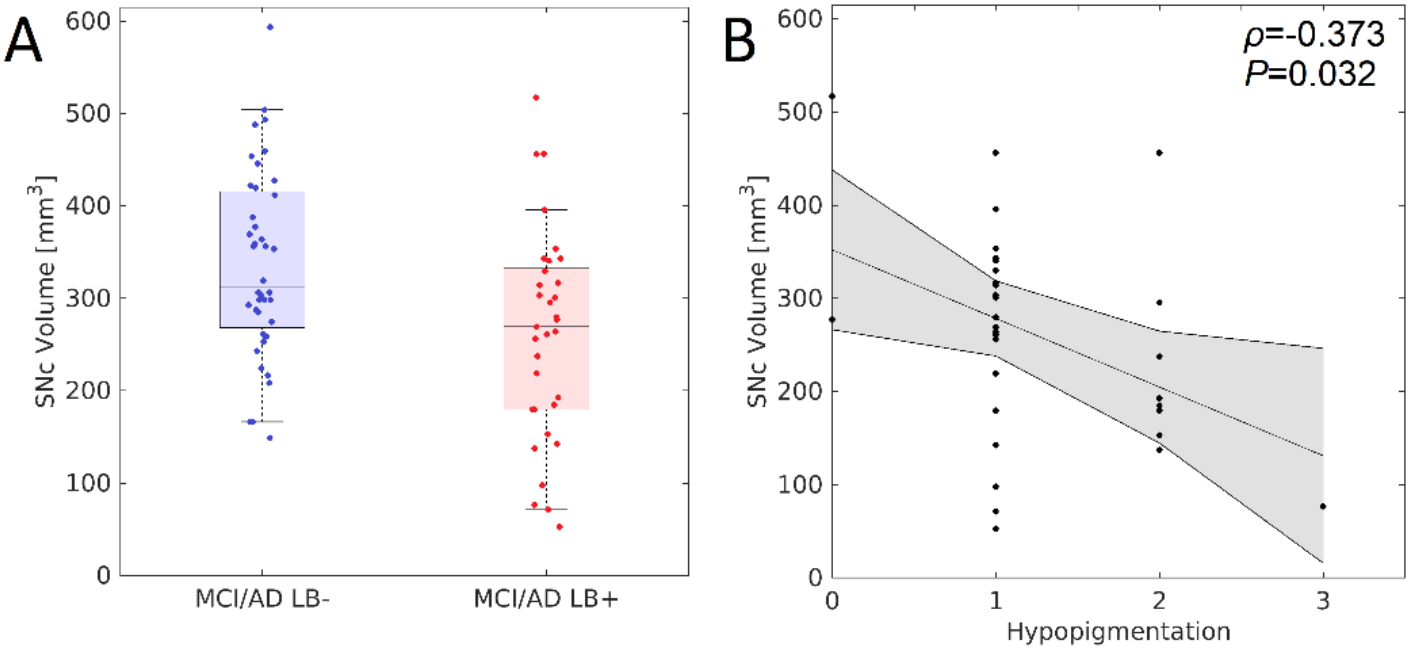
A comparison of SNc volume is shown in B and a significant reduction in nigral volume is seen in the AD/MCI LB+ group relative to the AD/MCI LB− group (A). A correlation between SNc volume and SNc neuronal loss is shown in (C) with greater neuronal loss correlating with reduced volume (B).

Within the LB+ group, lower SNc volume was associated with greater nigral hypopigmentation (Spearman ρ=−0.373; P=0.032). The association remained significant after adjusting for the imaging to autopsy interval (partial Pearson r=−0.418; P=0.019). Across leave one out analyses, the correlation between SNc volume and nigral hypopigmentation remained negative, with Spearman ρ ranging from −0.471 to −0.303. SNc volume was not significantly correlated with Trails B completion time, ADAS Q4 or ADAS11 scores, or UW executive function, visuospatial, or memory composite scores (Ps>0.070).

### 3.3 Lewy Body Status Discrimination

Logistic regression analyses included 71 participants (32 AD/MCI LB+ and 39 AD/MCI LB−) with complete data. Adding SNc volume to a model containing age, sex, and UW executive function composite score significantly improved the model fit for autopsy-defined LB status (likelihood ratio χ2(1)=8.67; P=0.003). The combined model yielded an AUC of 0.770 (95% confidence interval: 0.661-0.879). SNc volume and UW executive function composite score individually yielded AUCs of 0.679 (95% confidence interval: 0.574-0.820) and 0.691 (95% confidence interval: 0.569-0.812), respectively. These ROCs are plotted in Figure 3.

**Figure 3.**
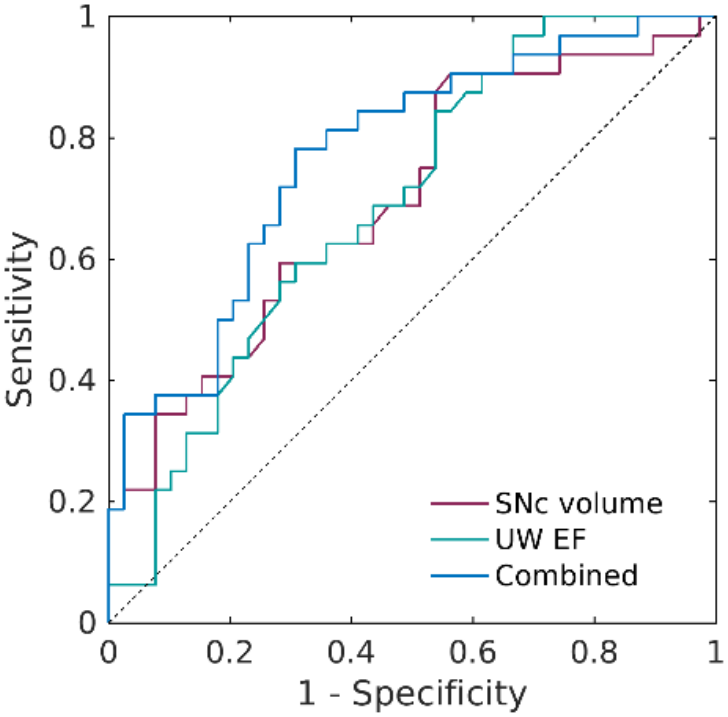
Receiver operator characteristic (ROC) curves for SNc volume, UW executive function (UW EF) composite score and the combined model.

## 4. Discussion

Although CSF-based SAAs enable antemortem detection of LB copathology ^16^, complementary imaging markers may help characterize neurodegeneration associated with LB copathology, inform participant stratification in AD clinical trials, and identify patients at risk of a more aggressive disease course ^9-12^. AD and MCI patients with autopsy confirmed LB copathology had greater nigral hypopigmentation and lower MRI-derived SNc volume than those without LB pathology. Within the LB+ group, lower SNc volume was associated with greater nigral hypopigmentation at autopsy, and this association persisted after adjustment for the imaging to autopsy interval. SNc volume also improved model fit for LB status beyond age, sex, and executive function.

Neuromelanin-sensitive contrast spatially colocalizes with melanized neurons in the SNc ^35^ and locus coeruleus ^36^. Other postmortem work has reported a correlation between neuromelanin-sensitive contrast and nigral neuromelanin concentration ^37^. Here, lower antemortem SNc volume was associated with greater nigral hypopigmentation at autopsy within the AD/MCI LB+ group even after adjusting for the imaging to autopsy interval. These findings provide additional evidence that neuromelanin-sensitive MRI reflects nigral pigmentation.

Postmortem studies have reported greater nigral neuronal loss and depigmentation in AD patients with LB copathology than in those with pure AD pathology ^10,17^. Consistent with these findings, AD/MCI LB+ patients in our cohort had higher nigral hypopigmentation and reduced nigral volume relative to AD/MCI LB− patients. Other studies have reported increased striatal FDG-PET uptake ^38^ and reduced nigral FDG-PET uptake ^39^ in AD/MCI LB+ patients relative to those without LB pathology. Taken together, structural and metabolic findings suggest that LB copathology is associated with alterations in the nigrostriatal system.

LB copathology in AD is associated with greater executive and visuospatial dysfunction ^9,40^, although these differences have not been consistently observed across studies ^14^. In the ADNI cohort, AD/MCI LB+ patients performed worse on executive function and visuospatial measures than AD/MCI LB− patients, despite no significant group difference in Braak stage. Despite these differences, executive function along provided modest discrimination of LB status (AUC = 0.691), consistent with the difficulty of identifying LB copathology through cognitive assessment, particularly early in the disease course ^15^. Adding SNc volume to age, sex, and executive function improved model fit (AUC = 0.770). These results suggest that SNc volume provides information about LB status beyond cognitive assessment.

Among participants with determinate SAA results, SAA classification was concordant with autopsy defined LB status in 41 of 45 cases (91.1%). This is consistent with published findings in the broader ADNI cohort ^16^. Antemortem identification of LB copathology using SAA could facilitate longitudinal studies of nigral integrity in AD and MCI patients. Serial neuromelanin-sensitive and iron-sensitive MRI in SAA-positive patients could help establish when nigral changes emerge and how they evolve over the disease course.

Several limitations should be considered when interpreting these findings. The dual echo TSE acquisition was not optimized for neuromelanin imaging and provides lower neuromelanin-sensitive contrast and poorer reproducibility than dedicated sequences with explicit MT preparation pulses ^41,42^, potentially reducing sensitivity to subtle nigral changes. Second, imaging preceded autopsy by an average of 5 years for AD/MCI LB− and 4 years for AD/MCI LB+, during which additional neurodegeneration may have occurred. Consequently, postmortem hypopigmentation may not fully reflect nigral integrity at the time of imaging. Although the volume hypopigmentation association persisted after adjustment for this interval, adjustment cannot account for individual differences in disease progression. Nigral hypopigmentation was assessed using a semiquantitative rating rather than quantitative measurements of neuromelanin content or melanized neuron density, limiting the precision of the MRI-pathology comparison. Finally, these findings are based on a single ADNI cohort. Replication in an independent cohort with both antemortem neuromelanin-sensitive and postmortem ratings of nigral hypopigmentation would strengthen the evidence of this association.

In AD and MCI patients, autopsy-confirmed LB copathology was associated with lower SNc volume, which also corresponded to greater nigral hypopigmentation within the LB+ group. SNc volume provided additional information about LB status beyond age, sex, and executive function. These findings support neuromelanin-sensitive MRI as a potential marker of nigral degeneration associated with LB copathology in AD.

## Data Availability

The ADNI data used in this manuscript are publicly available from the ADNI database (https://adni.loni.usc.edu) upon registration and compliance with the data use agreement.

## Acknowledgements

Data collection and sharing for this project was funded by the Alzheimer’s Disease Neuroimaging Initiative (ADNI) (National Institutes of Health Grant U01 AG024904) and DOD ADNI (Department of Defense award number W81XWH-12-2-0012). ADNI is funded by the National Institute on Aging, the National Institute of Biomedical Imaging and Bioengineering, and through generous contributions from the following: AbbVie, Alzheimer’s Association; Alzheimer’s Drug Discovery Foundation; Araclon Biotech; BioClinica, Inc.; Biogen; Bristol-Myers Squibb Company; CereSpir, Inc.; Cogstate; Eisai Inc.; Elan Pharmaceuticals, Inc.; Eli Lilly and Company; EuroImmun; F. Hoffmann-La Roche Ltd and its affiliated company Genentech, Inc.; Fujirebio; GE Healthcare; IXICO Ltd.; Janssen Alzheimer Immunotherapy Research & Development, LLC.; Johnson & Johnson Pharmaceutical Research & Development LLC.; Lumosity; Lundbeck; Merck & Co., Inc.; Meso Scale Diagnostics, LLC.; NeuroRx Research; Neurotrack Technologies; Novartis Pharmaceuticals Corporation; Pfizer Inc.; Piramal Imaging; Servier; Takeda Pharmaceutical Company; and Transition Therapeutics. The Canadian Institutes of Health Research is providing funds to support ADNI clinical sites in Canada. Private sector contributions are facilitated by the Foundation for the National Institutes of Health (www.fnih.org). The grantee organization is the Northern California Institute for Research and Education, and the study is coordinated by the Alzheimer’s Therapeutic Research Institute at the University of Southern California. ADNI data are disseminated by the Laboratory for Neuro Imaging at the University of Southern California.

## References

1. Brookmeyer, R., Johnson, E., Ziegler-Graham, K. & Arrighi, H.M. Forecasting the global burden of Alzheimer’s disease. Alzheimers Dement 3, 186–191 (2007).

2. Hardy, J. & Allsop, D. Amyloid deposition as the central event in the aetiology of Alzheimer’s disease. Trends Pharmacol Sci 12, 383–388 (1991).

3. Sayre, L.M., et al. In situ oxidative catalysis by neurofibrillary tangles and senile plaques in Alzheimer’s disease: a central role for bound transition metals. J Neurochem 74, 270–279 (2000).

4. Selkoe, D.J. The molecular pathology of Alzheimer’s disease. Neuron 6, 487–498 (1991).

5. Albert, M.S., et al. The diagnosis of mild cognitive impairment due to Alzheimer’s disease: recommendations from the National Institute on Aging-Alzheimer’s Association workgroups on diagnostic guidelines for Alzheimer’s disease. Alzheimers Dement 7, 270–279 (2011).

6. Rocca, W.A., et al. Trends in the incidence and prevalence of Alzheimer’s disease, dementia, and cognitive impairment in the United States. Alzheimers Dement 7, 80–93 (2011).

7. Lippa, C.F., et al. Lewy bodies contain altered alpha-synuclein in brains of many familial Alzheimer’s disease patients with mutations in presenilin and amyloid precursor protein genes. Am J Pathol 153, 1365–1370 (1998).

8. Hamilton, R.L. Lewy bodies in Alzheimer’s disease: a neuropathological review of 145 cases using alpha-synuclein immunohistochemistry. Brain Pathol 10, 378–384 (2000).

9. Azar, M., et al. Cognitive tests aid in clinical differentiation of Alzheimer’s disease versus Alzheimer’s disease with Lewy body disease: Evidence from a pathological study. Alzheimers Dement 16, 1173–1181 (2020).

10. Forstl, H., Burns, A., Luthert, P., Cairns, N. & Levy, R. The Lewy-body variant of Alzheimer’s disease. Clinical and pathological findings. Br J Psychiatry 162, 385–392 (1993).

11. Chung, E.J., et al. Clinical Features of Alzheimer Disease With and Without Lewy Bodies. JAMA Neurol 72, 789–796 (2015).

12. Mueller, C., et al. Survival time and differences between dementia with Lewy bodies and Alzheimer’s disease following diagnosis: A meta-analysis of longitudinal studies. Ageing Res Rev 50, 72–80 (2019).

13. Connor, D.J., et al. Cognitive profiles of autopsy-confirmed Lewy body variant vs pure Alzheimer disease. Arch Neurol 55, 994–1000 (1998).

14. Savica, R., et al. Lewy body pathology in Alzheimer’s disease: A clinicopathological prospective study. Acta Neurol Scand 139, 76–81 (2019).

15. Yoshizawa, H., Vonsattel, J.P. & Honig, L.S. Early neuropsychological discriminants for Lewy body disease: an autopsy series. J Neurol Neurosurg Psychiatry 84, 1326–1330 (2013).

16. Tosun, D., et al. A cross-sectional study of alpha-synuclein seed amplification assay in Alzheimer’s disease neuroimaging initiative: Prevalence and associations with Alzheimer’s disease biomarkers and cognitive function. Alzheimers Dement 20, 5114–5131 (2024).

17. Hansen, L., et al. The Lewy body variant of Alzheimer’s disease: a clinical and pathologic entity. Neurology 40, 1–8 (1990).

18. Sasaki, M., et al. Neuromelanin magnetic resonance imaging of locus ceruleus and substantia nigra in Parkinson’s disease. Neuroreport 17, 1215–1218 (2006).

19. Ogisu, K., et al. 3D neuromelanin-sensitive magnetic resonance imaging with semi-automated volume measurement of the substantia nigra pars compacta for diagnosis of Parkinson’s disease. Neuroradiology 55, 719–724 (2013).

20. Chen, X., et al. Simultaneous imaging of locus coeruleus and substantia nigra with a quantitative neuromelanin MRI approach. Magn Reson Imaging 32, 1301–1306 (2014).

21. Langley, J., et al. A multicontrast approach for comprehensive imaging of substantia nigra. Neuroimage 112, 7–13 (2015).

22. Castellanos, G., et al. Automated Neuromelanin Imaging as a Diagnostic Biomarker for Parkinson’s Disease. Mov Disord 30, 945–952 (2015).

23. Langley, J., Hwang, K.S., Hu, X.P. & Huddleston, D.E. Nigral volumetric and microstructural measures in individuals with scans without evidence of dopaminergic deficit. Front Neurosci 16, 1048945 (2022).

24. Hwang, K.S., Langley, J., Tripathi, R., Hu, X.P. & Huddleston, D.E. In vivo detection of substantia nigra and locus coeruleus volume loss in Parkinson’s disease using neuromelanin-sensitive MRI: Replication in two cohorts. PLoS One 18, e0282684 (2023).

25. Biondetti, E., et al. The spatiotemporal changes in dopamine, neuromelanin and iron characterizing Parkinson’s disease. Brain 144, 3114–3125 (2021).

26. Vitali, P., et al. Substantia Nigra Volumetry with 3-T MRI in De Novo and Advanced Parkinson Disease. Radiology 296, 401–410 (2020).

27. Schwarz, S.T., Xing, Y., Tomar, P., Bajaj, N. & Auer, D.P. In Vivo Assessment of Brainstem Depigmentation in Parkinson Disease: Potential as a Severity Marker for Multicenter Studies. Radiology, 160662 (2016).

28. Takahashi, H., et al. Imaging of the nigrostriatal system for evaluating the preclinical phase of Parkinson’s disease development: the utility of neuromelanin, diffusion MRI, and DAT-SPECT. Br J Radiol 95, 20210837 (2022).

29. Langley, J., Hwang, K.S., Huddleston, D.E., Hu, X.P. & Parkinson’s Progression Marker, I. Nigral volume loss in prodromal, early, and moderate Parkinson’s disease. NPJ Parkinsons Dis 11, 181 (2025).

30. Inagawa, Y., et al. Utility of neuromelanin-sensitive MRI in the diagnosis of dementia with Lewy bodies. PLoS One 19, e0309885 (2024).

31. Dixon, W.T., Engels, H., Castillo, M. & Sardashti, M. Incidental magnetization transfer contrast in standard multislice imaging. Magn Reson Imaging 8, 417–422 (1990).

32. Smith, S.M., et al. Advances in functional and structural MR image analysis and implementation as FSL. NeuroImage 23 Suppl 1, S208–219 (2004).

33. Woolrich, M.W., et al. Bayesian analysis of neuroimaging data in FSL. NeuroImage 45, S173–186 (2009).

34. Langley, J., et al. Multimodal assessment of nigrosomal degeneration in Parkinson’s disease. Parkinsonism & related disorders 80, 102–107 (2020).

35. Kitao, S., et al. Correlation between pathology and neuromelanin MR imaging in Parkinson’s disease and dementia with Lewy bodies. Neuroradiology 55, 947–953 (2013).

36. Keren, N.I., et al. Histologic validation of locus coeruleus MRI contrast in post-mortem tissue. Neuroimage 113, 235–245 (2015).

37. Cassidy, C.M., et al. Neuromelanin-sensitive MRI as a noninvasive proxy measure of dopamine function in the human brain. Proc Natl Acad Sci U S A 116, 5108–5117 (2019).

38. Kantarci, K., et al. FDG PET metabolic signatures distinguishing prodromal DLB and prodromal AD. Neuroimage Clin 31, 102754 (2021).

39. Lee, Y.H., et al. Effect of Alzheimer’s Disease and Lewy Body Disease on Metabolic Changes. J Alzheimers Dis 79, 1471–1487 (2021).

40. Quadalti, C., et al. Clinical effects of Lewy body pathology in cognitively impaired individuals. Nat Med 29, 1964–1970 (2023).

41. Wengler, K., He, X., Abi-Dargham, A. & Horga, G. Reproducibility assessment of neuromelanin-sensitive magnetic resonance imaging protocols for region-of-interest and voxelwise analyses. Neuroimage 208, 116457 (2020).

42. van der Pluijm, M., et al. Reliability and Reproducibility of Neuromelanin-Sensitive Imaging of the Substantia Nigra: A Comparison of Three Different Sequences. J Magn Reson Imaging 53, 712–721 (2021).

